# Subjective survival probabilities by employment category and job satisfaction among the fifty-plus population in Japan

**DOI:** 10.1101/2023.01.01.23284103

**Authors:** Jacques Wels

## Abstract

**Background:** Subjective Survival Probabilities (SSP) are known to be associated with mortality but little is known about the relationship they might have with employment categories and job satisfaction. We assess such a relationship looking at the fifty-plus population in Japan that is characterized by a stratified labour market for the older workers and high working time intensity.

**Method:** We use the four waves (2007-2013) of the Japanese Study of Aging and Retirement (JSTAR), a panel dataset tracking 7,082 50-plus respondents in 10 Japanese prefectures. We use a mixed-effects quantile regression model to investigate the relationship between SSP and employment status (model 1) and job satisfaction (model 2). Both models additively control for demographic and socio-economic cofounders as well as other health measurements. Multiple imputations are used to correct sample attrition.

**Results:** In model 1, retirement (−0.27, 95%CI =-0.51;-0.03) and contract work (−0.51, 95%CI=-0.79;-0.23) are negatively associated with SSP in comparison with full-time employment. In model 2, low job satisfaction appears to be strongly associated with SSP (−1.37, 95%CI=-1.84;-0.91) in comparison with high job satisfaction. The same trend is observed regardless of the way job satisfaction is calculated. Both working time and employment category are not significantly associated with SSP after controlling for job satisfaction which indicates that job satisfaction is a main driver of SSP discrepancies.

**Discussion:** SSP variations can be explained by employment category with contract work more at risk. Job dissatisfaction is a main explanation of low SSP. Both work and employment explain SSP variations.

## Background

Japan has one of the highest life expectancy in the world with a life expectancy at birth of 84.7 in 2020 (a record of 87.7 for women and 81.6 for men), just after Norway (81.7), Iceland (81.8) and Switzerland (81.9) (OECD, 2021). Life expectancy is an opportunity and a challenge and the multiple pension reforms that have been implemented in Japan throughout the past two decades partially reflect the unbalance between an ageing population and a constant drop in natality (Wels & Takami, 2021). Against such a backdrop, little is known on the way individuals perceive their own life expectancy and the work and employment factors that could explain discrepancies in perceiving it.

Subjective Survival Probabilities (SSP) – also called subjective life expectancy or self-rated life expectancy – as well as self-rated health are both strongly associated with mortality although much more is known about self-rated health than about measures of self-predicted life expectancy that only emerged in the early 2000 (Jylha, 2011). Although, there is evidence that differences exist across the two measurements. For instance, smoking is associated with lower subjective survival probabilities but not lower self-rated health. By comparison, arthritis is associated with lower self-rated health but does not affect subjective life expectancy (Jylha, 2011). All types of chronic conditions do not affect SSP the same way (Vanajan & Gherdan, 2022). SSP appears to be a direct indicator of the probability of mortality that is not framed by self-rated health, even though self-rated health could predict mortality (Cho et al., 2022; Idler & Benyamini, 1997; Mossey & Shapiro, 1982). Similarly, health behaviours are seen as dominant attributes of self-rated health whilst family longevity history, life situations and lack of control may impact SSP (Lee et al., 2020). SSP is constructed based on the understanding of one’s personal life expectancy based on factors that are similar to those predicting actual life expectancy but with differences in the way some factors are weighted. Therefore, SSP is considered as a powerful predictor of mortality after adjusting for self-rated health (Kim & Kim, 2017). Gender differences exist when looking at SSP. The effect of SSP on mortality is stronger for men than women (van Doom & Kasl, 1998) suggesting a possible under estimation of life expectancy among female.

However, few studies have paid attention to the socio-economic determinants of SSP that play a major role in explaining discrepancies across populations (Bae et al., 2017). Using UK panel data, it was shown that SSP is strongly associated with lifetime socio-economic position (i.e., social class and household incomes) (Popham & Mitchell, 2007). Authors also confirm that, after controlling for self-rated health and limiting illness, SSP is partially independent of current health status. SSP is particularly important in late career and near the so-called retirement zone (Vickerstaff, 2006) as subjective life expectancy is a factor that is considered by workers in the retirement decision making as employees with longer predicted life expectancy tend to wish to retire later but this not translate into effects and no difference is observed in terms of retirement behaviours (van Solinge & Henkens, 2018).

In this study, we specifically look at the relationship between SSP and work and employment within the population aged 50 and over in Japan. Employment is understood as a status within or outside the labour market that can be associated with in-kind – such as earnings or pensions benefits – or in-nature benefits – such as a status within the society – whilst work refers to the task that is actually done and could be associated with a status, or not (Jahoda, 1982).

To operationalize these two notions, we first focus on the different possible status the 50+ population could occupy. One of the main issues Japanese policy makers have faced in their attempt to increase effective ages of retirement and employment participation among the elderly is about the gap between the increasing pension eligibility age and the mandatory retirement age that is fixed to 60 in most companies (Kondo & Shigeoka, 2017). The Japanese Government has gradually pushed for increasing retirement ages and employment participation among the elderly through two main reforms: The Pension Reform Act (PEA) and the Elderly Employment Stabilization Law (EESL). On the one hand, the PEA was implemented in 2001 to gradually raise the eligibility age from 60 to 65 for the fixed part of the pension benefit (i.e., the part that is not proportional to past earning but common for all retired) (Kondo & Shigeoka, 2017) together with a declining generosity in calculating pension benefits (Oshio et al., 2018). On the other hand, the EESL was revised in 2006 and has compelled companies to either increase their retirement age to make it equal to the pension eligibility age, to clarify formal rules for employment extension or reemployment or to abolish the mandatory retirement age. The EESL was revised again in 2013 with the abolishment of the selection of employees based on certain criteria. The main objective of the EESL is to increase retention rate for those already employed but with no direct impact on the elderly’s incentives to work (Oshio et al., 2018). For the comparison, one main specificities of the EESL in Japan compared with anti-age discrimination laws in the US, the UK or Canada is that the EESL explicitly targets the protection of those just before the pension eligibility age (Kondo & Shigeoka, 2013). The gap between pension eligibility and company mandatory retirement ages has led many companies to adopt a reemployment scheme instead of increasing their retirement age to 65. The common reemployment scheme is to have employees retire once at retirement age set by the company and sign a reemployment contract in a non-regular employment form as a contract employee (*shokutaku*). The health implications of these forms of late career contract work have not been fully understood yet but studies suggest a negative long-term relationship with self-reported health (Wels, 2019b; Wels & Takami, 2021) and indirect effects such as job transfer (e.g., to another branch) or resignation (Jiang, 2021). Contract work is a main feature of late career transitions in Japan, but the retirement zone is also characterized by other statuses. On the one hand, many workers are not employed in Japan. A substantial part of the workforce if self-employed or working in an independent business and, among the female population (particularly among the oldest cohorts) a large part does domestic work. On the other hand, part-time employment exists but no regulation was implemented to support gradual retirement as it is the case in other countries (Wels, 2019a).

The second aspect taken into consideration in this study is work, and more specifically working time and job satisfaction that are both detrimental in Japan. Despite having fallen below the US in recent years, average working time in Japan remains one of the highest within the OECD (Takami, 2019). There is a well-known relationship between job satisfaction and self-reported health. Job satisfaction is seen as one of the factors having the highest influence on workers’ health and reflects the type of work that people do (Bartley et al., 2005) but also the quality of the employment that they have (van Aerden et al., 2016). Several factors play a role in explaining variations in job satisfaction within the workforce. First, scheduling control and work-life balance policies are connected to job satisfaction (Jang et al., 2011). Second, perceived age discrimination among the older workforce is also associated with lower job satisfaction whilst support from supervisors and co-workers reduce such a negative impact (Harada et al., 2019; Yamaguchi, 2013). Thirdly, earnings could play a role in explaining job satisfaction but with different degrees of intensity depending on the country with a greater impact of non-financial aspects in East Asia (Yeh, 2015), particularly for the older workforce where job satisfaction seems more driven by relationships with colleagues than by job security, advancement opportunities or job security (Iwasai et al., 2006). Finally, working time also interacts with job satisfaction in explaining workers’ health but, in Japan, long working hours only have a negative effect on health for those who report a low job satisfaction highlighting that job satisfaction improvements could reduce the negative relationship between working time and poor health (Nakata, 2017). In other words, the relationship between overtime and job overload is less pronounced than the one with job control, skill use and support from colleagues (Kawakamii & Haratani, 1999). For instance, a recent study suggests that working time reduction in late career in Japan is not associated with mental health improvements (Minami et al., 2015).

This short review shows some knowledge gaps that remain to be investigated. First, there is often a confusion between work and employment, and job satisfaction may be – depending on the study – affected by the employment category people falls in. Distinguishing the nature of the task, social support, pay, on the one hand, and the status that is associated with one’s position within the labour market seems important because they refer to two different areas of policy intervention. Second, job satisfaction is not properly defined. Some studies rely an overall self-reported job satisfaction variable whilst some other focus on some specific aspects such as pay adequacy, physical work or colleagues’ support. Combining different measurements of job satisfaction – all subjective but focusing on different aspects – could be of interest. Finally, little is known on the relationship between employment categories, job satisfaction and subjective survival probabilities. Most studies have focused on self-reported health, mental health, or health conditions but the specificities of SSP have not been assessed when looking at work and employment, particularly in Japan where work intensity is high and employment categories for the older workforce are specific.

Using panel data collected over four waves between 2007 and 2013 within ten Japanese prefectures, this study assesses the employment and work factors associated subjective survival probabilities discrepancies. We specifically address four research question: (RQ1) Is there a difference in SSP by employment status among the full 50+ population? ; (RQ2) Does SSP vary by job satisfaction and working time among the employed and self-employed population? ; (RQ3) What is the specific relationship of job satisfaction and employment category on SSP?; (RQ4) Do adjustments by demographic, socio-economic and objective and self-reported health cofounders affect the nature these relationships?

## Data and methods

### JSTAR

Data come from the Japanese Study of Aging and Retirement (JSTAR) that is a longitudinal dataset that currently contains four waves collected in 2007, 2009, 2011 and 2013. The original sample strategy is described in (Ichimura, Shimizutani, & Hashimoto, 2009). At the baseline (2007), JSTAR includes respondents aged 50 to 75 living in five municipalities in eastern area of Japan (Ichimura et al., 2009): Takikawa in Hokkaido, Sendai in the Tohoku area, Adachi Ward within Tokyo, Kanazawa in Hokuriki and Shirakawa in the Chubu area. Some efforts have been made to increase the number of cities. The 2009 wave includes a refreshment sample from two additional cities (Tosu and Naha) and the 2011 wave includes another refreshment sample from three additional cities (Chofu, Hiroshima and Tondabayashi). In total, the sample focuses on ten cities. Sample design is shown in <u>supplementary file 1</u>. The full sample includes 16,932 observations over four time-points including a total of 7,082 respondents.

### Subjective Survival Probabilities (SSP)

JSTAR contains repeated questions on life expectancy where it is asked to the respondents the probability (in percentage) they believe they are going to live at least until some specific ages (75, 80, 85, 90, 95, 100, 105, 110). Unexpectedly, due to the age nature of the sample, the self-predicted probabilities to live under 70 and 75 are high and the probabilities to live beyond 95 are extremely low but the distribution is sparser when looking at ages 80, 85, 90 and 95, which corresponds to the life expectancy at birth reported by the OECD (OECD, 2021). However, even if calculated as percentages, probabilities at not normally distributed for the three age limits with peaks at 0, 10, 20, 50, 70, 80 and 100 percent and highest density in the highest probabilities at age 80 and at the lowest probabilities at age 90.

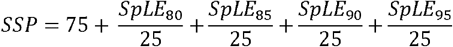

To tackle this issue and provide a single indicator of SSP, we have generated a normally distributed variable that combines information collected for four age-limits (80, 85, 90 and 95), i.e., the self-predicted probability that respondents live beyond these specific age bands. We divide each age limit probability by 25 for them to spread over a four-year period and add up each probability to the original threshold of 75 (the lower age-limit information was collected on).

### Employment categories

The employment category variable combines information flowing from three variables: (1) whether respondents were working (including in temporary leave) at the time of the survey; (2) if working, the type of work they were doing including employee, company executive, contract employed, temporary employed, owner of an independent business, helping in an independent business or doing a side job at home; finally, (3) for those employed (the majority of the sample after those not working), we use information on their working time pattern to distinguish full-time working, part-time workers and other types of work patterns. This results in ten different categories. ‘Not working’ being the most common status among the sample, we divide this category into four sub-categories that include information on the reasons for not working including being retired, keeping house, receiving medical care or any other reasons (including unemployment). Consequently, the employment status contains 13 categories: (1) employed full-time, (2) company executive, (3) employed part-time, (4) employed under contract, (5) temporary employed, (6) employed other, (7) owner of independent business, (8) help in independent business, (9) side job at home, (10) retired, (11) not retired – receiving medical care, (12) not retired – keeping house, (13) not retired – other reasons. The first modality (employed full-time) is selected as the reference category.

### Job satisfaction and working time

Among the working population, we look at several indicators of job satisfaction. The first indicator that is used is a self-reported variable on overall job satisfaction (‘Overall, I am satisfied with my current job’). The variable contains four modalities: strongly agree, somewhat agree, do not really agree and strongly disagree. The first category is selected as the reference. To ensure that the self-reported job satisfaction accurately reflects different aspects of work, we run sensitivity analyses using a numeric variable accounting for seven levels of job satisfaction including whether the job include physical labour, whether respondents have a lot of work and feel time pressure, whether respondents have discretion about how they do their work, whether they receive help and support from colleagues, whether they receive appropriate evaluation of their work by co-workers, whether they are satisfied with their pay and whether they believe they are at risk of losing their job for other reasons than retirement. We sum up values for each variable and divide the final value by 7. Other variables include total working time including overtime (less than 20 hours/week, 21 to 30 hours/week, 31 to 40 hours/week, 41 to 50 hours/week, 51 to 60 hours/week, more than 60 hours/week), working time pattern (generally same hours; hours vary each week; seasonal work) and total number of holidays including bank holidays

### Covariates and Adjustment levels

The model controls for the following covariates: *Age* (in years of age); *Gender* (female, male – male being the reference category) ; *Highest level of education* obtained (distinguishing elementary to middle school, high school, junior college, vocational school and university degree – the latter being the reference category); *Marital status* (distinguishing those who are married or have a common law spouse versus those who are not); Whether the *household borrowed money* to friends or family (‘yes’, ‘no’ – ‘no’ being the reference category); Whether the household *rent* their accommodation (‘yes’, ‘no’ – ‘no’ being the reference category); Whether the respondent has a *private health insurance* under their own name (‘yes’, ‘no’ – ‘no’ being the reference category); *Self-reported health* (coded on a 5-item scale ranging from ‘good’ to ‘Not good and used as a numeric variable); *Multimorbidity* (distinguishing respondents who declare having been diagnosed with at least two health conditions including heart disease, high blood pressure, hyperlipemia, cerebral accident, diabetes, chronic lung disease, liver disease, ulcer or stomach disorder, joint disorder, bladder disorder, depression or emotional disorder, cancer); General Health Questionnaire (*GHQ-20*) caseness that is based on the answers to twentu items (from 1. Not at all to 5. 5 days a week or more) that are summed up and categorized into two categories across the time-point means (‘yes’ or ‘no’, ‘no’ being the reference category); Whether the respondent was an *outpatient* at an hospital over the past year (‘yes’ or ‘no’, ‘no’ being the reference category); Whether the respondent spent at least *one night at the hospital* over the past year (‘yes’ or ‘no’, ‘no’ being the reference category); *life satisfaction* that distinguishes those reporting being somewhat unsatisfied or unsatisfied and those reporting being satisfied or very satisfied (reference category). All the variables used in this study are time-varying except gender, highest level of education, marital status and whether the respondent has a health insurance that are fixed.

To better interpret the effect of these variables, the models includes three layers of adjustment: (1) The *unadjusted model* only controls for gender and age. (2) The *socio-demographic adjusted* model additively controls for the highest level of education obtained, the marital status, whether the household borrowed money to friends or family, whether the household rent or own its accommodation and whether the respondents has a private health insurance.(3) The *fully adjusted model* additively controls health variables including self-reported health, whether the respondent was diagnosed with two or more health conditions (multimorbidity), GHQ-20, whether the respondent was an outpatient at an hospital over the past year, whether the respondent spent the night at the hospital, life satisfaction,

### Models

We use linear quantile mixed models (LQMM) based on asymmetric Laplace distribution (Geraci & Bottai, 2007) to avoid bias due to the distribution of SSP. Unlike the linear regression that aims to explain the mean values of a scale outcome, the quantile regression looks at specific percentiles, in this case the median. The mixed model accounts for the four time points included in the dataset as well as for individual levels. Results flowing from the LQMM are compared with results flowing from a traditional linear mixed model regression. Analyses are performed on R using the packages ‘lqmm’ (Geraci, 2014) and the command ‘lmer’ from the package ‘lme4’ (Bates et al., 2014). We use two different modelling. Model 1 focuses on employment status for the full population (i.e., 16,932 observations over four time-points – 7,082 respondents). Model 2 focuses on job satisfaction and employment status among the working population (i.e., 8,689 observations for four time-points – 3,898 respondents).

### Missing data

Given the sample characteristics, repeated measurement is likely to lead to high attrition rates. We assess data missingness across waves and apply multiple imputations to correct sample bias due to attrition. We then meta-analyse the estimates flowing from the subsequent imputed datasets. We also compare results flowing from the non-imputed models with results flowing from the imputed model.

## Results

### Descriptive statistics

Descriptive statistics on the variables used in this study for the full-observations and by time point are shown in <u>supplementary file 2</u>.

Figure 1 shows a density plot of SSP by gender (female, male) and time point (2007, 2009, 2011 and 2013) as well as the overall mean by gender among the four time points. It can be observed that the mean for female is higher than for male with 83.8 years of age against 83 for men, a difference of 0.8 years of age that does not reflect the actual difference shown by the life expectancy at birth that is of 5.9 years of age. Calculating the median age by gender further narrows the gap with a median of 83.4 for women and 83 for men as it reduces the weight of extreme values among the female sample. Another interesting aspect of figure 1 is that it shows change across time points. One observes that the younger are the respondents, the more likely they are to report a low predicted life expectancy. This is the result of two combined factors. On the one hand, the sample composition is made of refreshment samples in 2009 and 2011 that reduce the average age of the sample. On the other hand, one could assume that being younger could be associated with a less foreseeable future. In other words, as age increase, SSP to live until old age increase as well.

**Figure 1.**
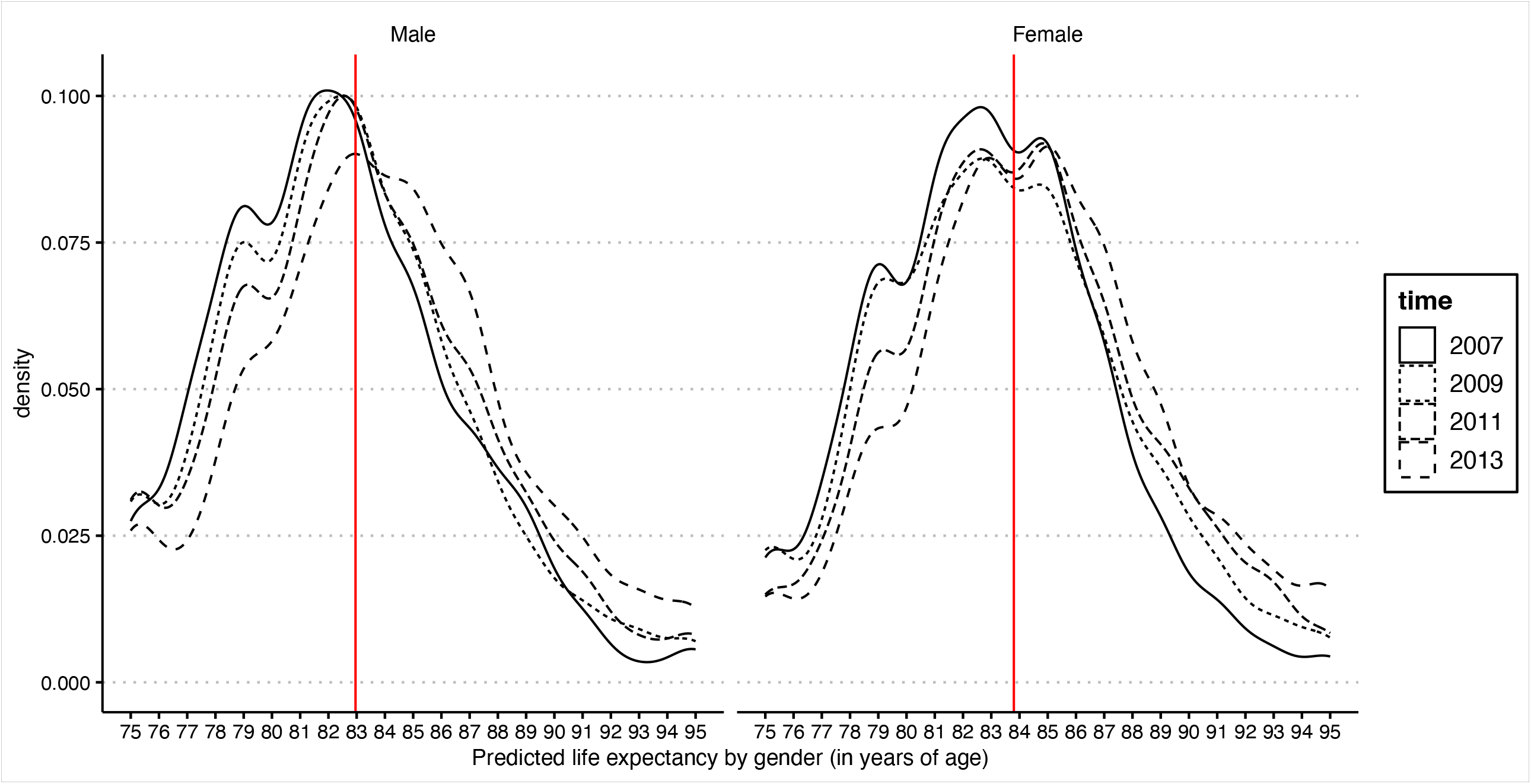
Density plot of self-predicted life expectancy by gender and wave

This study is interested in how the employment status and job satisfaction could play a role in explaining SSP among the population. Figure 2 shows the density plot of predicted life expectancy by job satisfaction (as a factor variable) among the four waves as well as the mean by category. The mean for those very highly satisfied is 83.6 years of age against, respectively, 83.2, 82.5 and 81.2 for those highly, low or very low satisfied. Similarly, the median is 83 for those very highly satisfied against, respectively, 83, 82.3 and 81 years of age for the other groups. At a descriptive level and without controlling for other variables, there is a clear relationship between both variables.

**Figure 2.**
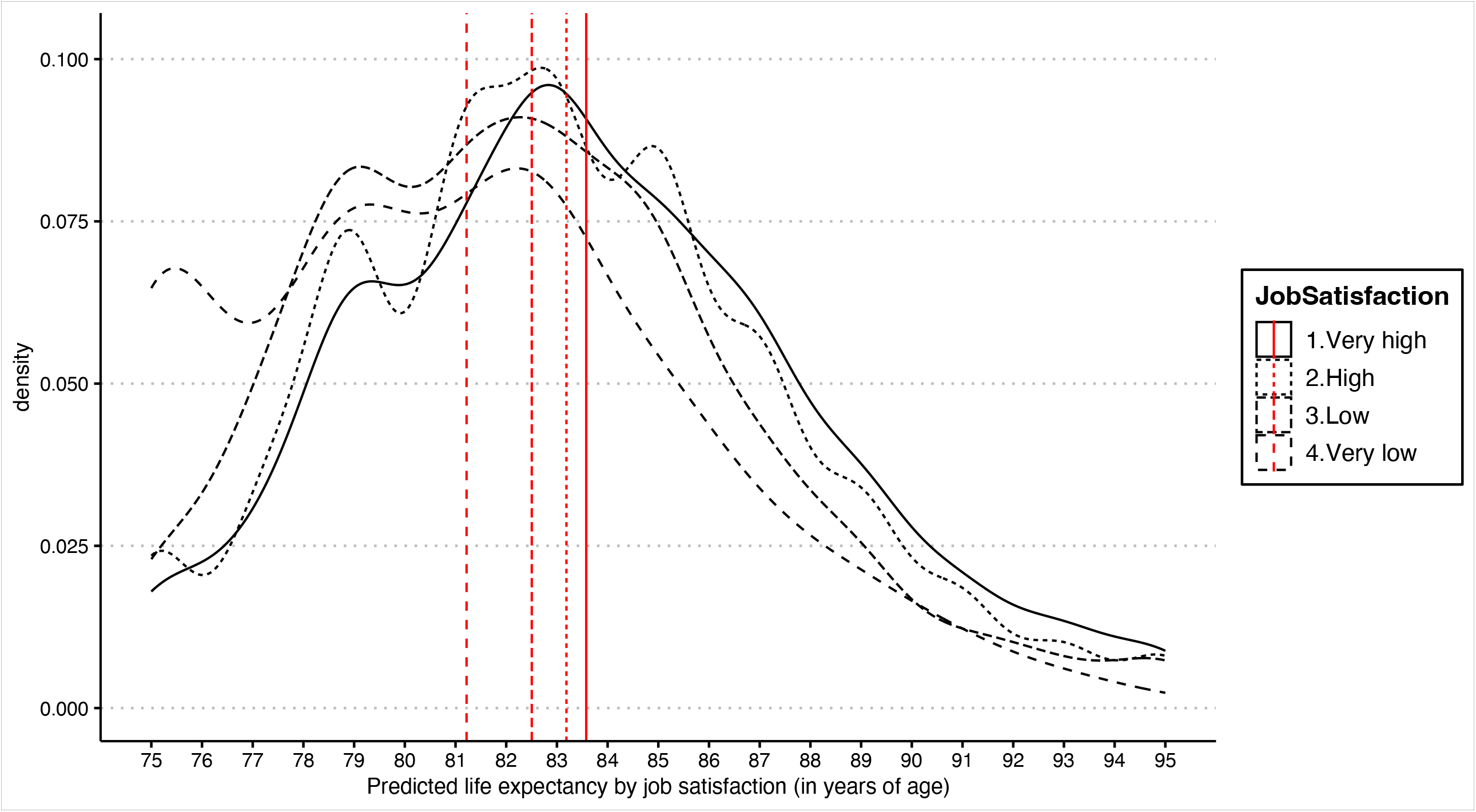
Density of self-predicted life expectancy by job satisfaction

The same type of relationship is observed when looking at the employment status (see <u>supplementary file 3</u>) where the SSP mean greatly varies from, for instance, 82.5 years old for the full-time employed population against 83.9 within the retired population or 82.2 for those reporting housekeeping as their main occupation. By comparison, those under contract work expect to leave until an average of 83. Median ages are also different with 82.2 years old for the full-time employed against 83.4 for the retired, 84.2 for the house keepers and 83 years old for the contract workers. Table 1 shows the representativeness of each employment category within the sample with full-time workers, contract employees, retired respondents and housekeepers representing respectively 14.7, 4.4, 15.6 and 25 per cent of the total observations

**Table 1.**
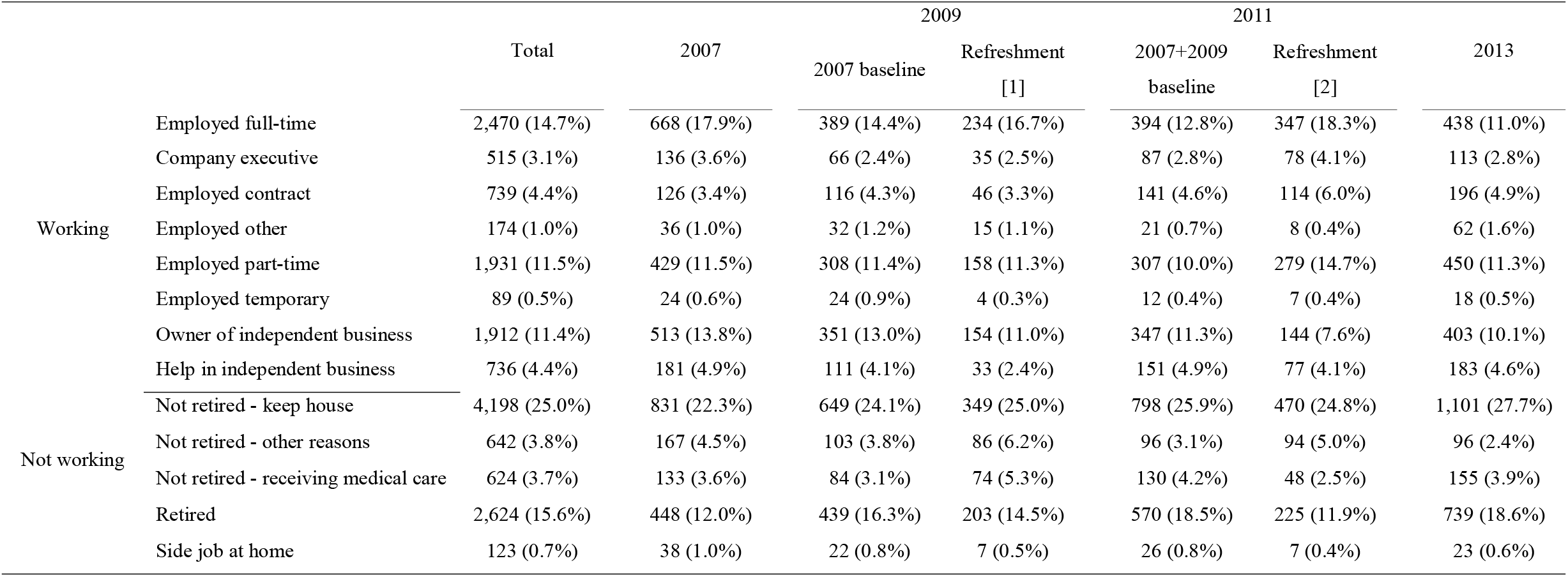
Employment categories by time point.

These descriptive figures poorly reflect the nature of SSP because they do not control for cofounding factors such as gender and age that are strong predictors of life expectancy. They also account for the sum of waves as if it were about different individuals whilst the sample is longitudinal. Finally, they show an association that is cross-sectional and not a potential change in association that would be observed over time.

### Relationship between employment category and SSP (Full population)

Figure 3 summarizes the main findings flowing from model 1 using a LQMM regression over three levels of adjustments and after multiple imputations. Full estimates and 95%CI from both the LQMM and the linear model after multiple imputations are shown in <u>supplementary file 4</u>.

**Figure 3.**
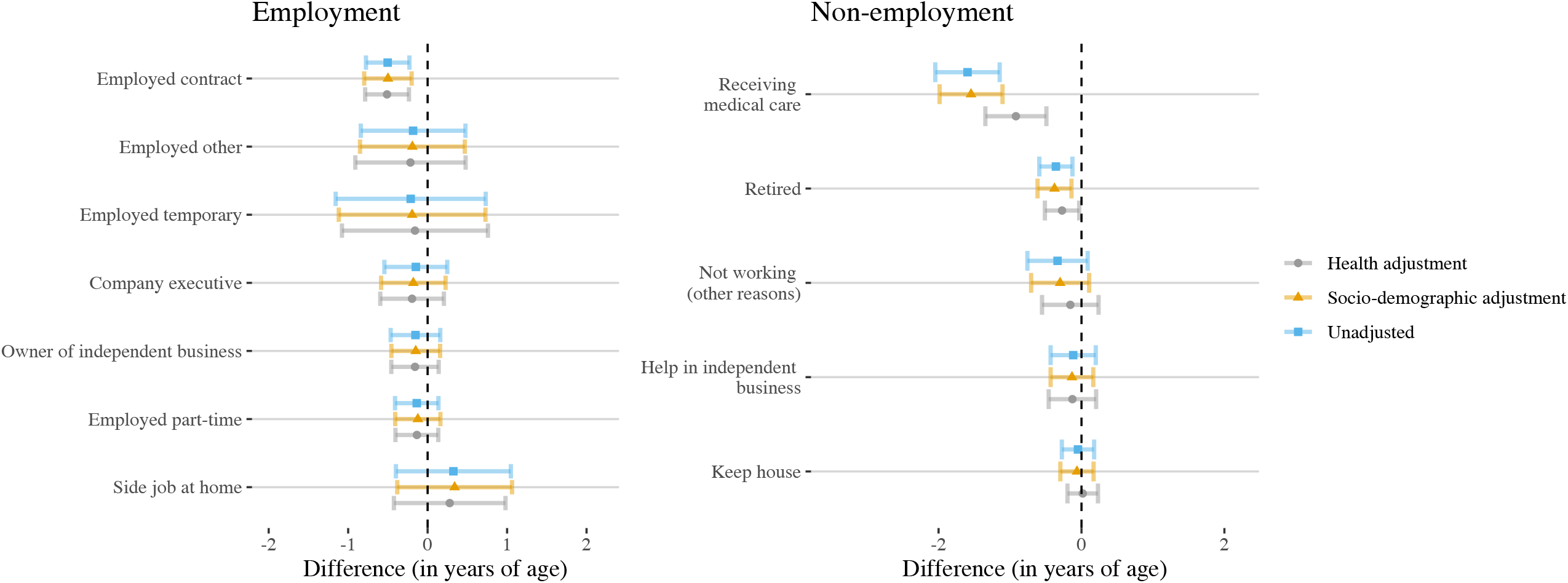
Model 1 – LQMM, main estimates

What figure 3 shows is that no employment category seems to do significantly better that full-time employment (that is the reference category). As expected, those receiving medical care report a SSP that is significantly lower compared to full-time employed by -1.60 years of age (95%CI: -2.05, -1.15) in the unadjusted model, -1.55 (95%CI: -1.99, -1.11) in the socio-demographic adjusted model and -0.92 (95%CI: -1.35, -0.49) in the health adjusted model. To a lesser extent, but still significatively, those employed under contract also report a much lower SpLE in comparison with full-time workers by, respectively, -0.50 (95%CI: -0.78, - 0.23), -0.50 (95%CI: -0.80, -0.20) and -0.51 (95%CI: -0.79, -0.24) years of age. A similar trend is observed for those who retired but with smaller differences: -0.36 (95%CI: -0.59, - 0.13), -0.38 (95%CI: -0.62, -0.14) and -0.27 (95%CI: -0.51, -0.03). Other estimates indicate a slight negative relationship for company executive, owner of independent business, part-time employed, not working for other reasons such as unemployment, helping in independent business and keeping house but not statistically significant at 95 percent.

Some cofounders play a major role in explaining SSP differences across the population such as age that is positively associated (between 0.13 and 0.14 depending on the adjustment) and gender (being a female compared to a men is associated with an increase in SSP median of between 0.43 and 0.46). The highest level of education obtained is associated with SSP in the socio-demographic adjusted model but with lower intensity (and significance) in the health adjusted model with university degree (reference category) associated with higher SSP. Not being married or having a common spouse is negatively associated with SSP, though estimates are not significant. As for education, renting an accommodation (instead of owning it) is associated with lower SSP in the socio-demographic adjusted model (−0.24, 95%CI: - 0.42, -0.06) but the relationship is smaller and not significant in the health adjusted model (−0.05, 95%CI:-0.22, 0.11). Finally, it has to be noted that all health indicators are independently associated with SSP including self-reported health, multimorbidity, GHQ caseness, having been an outpatient at a clinic or hospital or having spent at least one night at hospital over the past year, and life satisfaction.

### Relationship between job satisfaction and SSP (Working population)

The employment status relates to SSO with higher predicted ages across those in full-time employment compared with retirement or contract work. Model 2 focuses on those in employment only (removing those retired, keeping house or in medical care from the sample) and on the specific relationship job satisfaction could have with SSP after controlling for the employment status and other covariates. Figure 4 exhibits the main estimates flowing from model 2 using LQMM with employment categories on the left side of the figure and job satisfaction (as factor) on the right side. Full estimates from the LQMM and linear models are shown in <u>supplementary file 5.</u>

**Figure 4.**
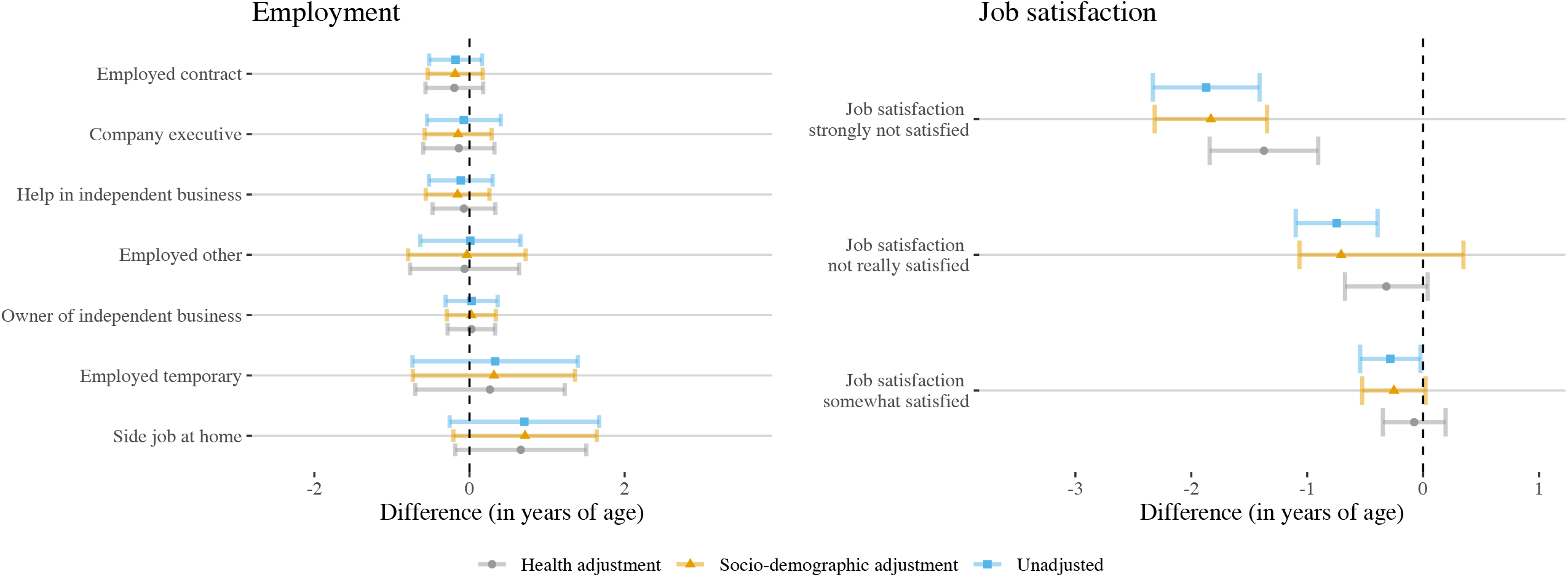
Model 2 – LQMM, main estimates

What figure 4 shows is that, after controlling for job satisfaction, the intensity of the coefficient for employment is lower and results are not statistically significant. We again observe negative relationships between SSP and contract work (−0.19, 95%CI: -0.54, 0.17 in the socio-demographic adjusted model) or helping in an independent business (−0.15, 95%CI: -0.57, 0.26) but these are attenuated. By contrast, job satisfaction plays an important role. In comparison with those being fully satisfied by their job, those somewhat satisfied report a SSP lower by 0.08 (95%CI: -0.35, 0.19) in the health adjusted model, -0.32 (95%CI: -0.67, 0.40) for those not really satisfied and -1.37 (95%CI: -1.84, -0.91) for those strongly not satisfied. Levels of adjustment tend to successively reduce the intensity of the coefficients with stronger estimates in the non-adjusted model and less stronger associations when controlling for health factors. One reason for this is that SSP is strongly associated with objective and subjective health measurement and controlling for these variables reduces the power of the model. Nevertheless, job satisfaction is more strongly associated with SSP, showing that, after controlling for employment statuses, job satisfaction is as much or even more relevant in understanding life expectancy predictions.

Analyses for model 2 were replicated using an item-based definition of job satisfaction instead of a self-reported unique question. Estimates using such a variable are shown in <u>supplementary file 6</u>. Results are not different from those observed above with a negative relationship between job satisfaction and SSP.

### Sensitivity checks

Estimates in the mixed effect linear and in the linear quantile mixed effects regression after multiple imputations are not significantly different in model 1 as can be observed in <u>supplementary file 4</u>. For instance, contract work is negatively associated with SSP in both models but with a slightly higher intensity in the quantile regression (−0.51 in the health adjusted model versus -0.42) and the relationship with retirement is also more pronounced within the quantile regression (−0.27 versus -0.25). The same conclusion can be made for model 2 (<u>supplementary file 5</u>) where coefficients are of the same nature but with slightly higher intensity in the quantile regression. Results from the complete case analysis (before multiple imputations) are shown in <u>supplementary files 7, 8 and 9</u> for models 1 and 2. It can be seen that multiple imputations does not drastically change the direction of the estimates (fully adjusted contract work is -0.11 in the linear model and -0.16 in the quantile model) but instead increase their intensity and reduce their significance levels. By contrast, using a complete case analysis with the item-based definition of job satisfaction increase the significance levels without changing the estimates, as can be observed in <u>supplementary file 9</u>.

## Discussion

Studies have focused on the determinants of subjective survival probabilities for more than twenty years. However, little is known on their relationship with employment participation and job satisfaction, particularly in the context of Japan where the labour market for the older workforce is stratified by gender and where specific employment arrangement exist to maintain the older workforce in employment. Using panel data for the population aged 50 and over, the main objective of this study was to assess the nature of such a relationship and to better understand how employment and job satisfaction could play a role in explaining SSP discrepancies.

This study is not without limitations. Two main limitations flow from the dataset. First, information on sector of activity was not consistently coded across waves which makes the inclusion of a sector variable difficult. Second, information on household income was not consistently collected across waves with different information on social benefits, extra-work benefits and household total earnings which also makes difficult the use of such a variable. To tackle this bias, we have used several socio-economic variables such as education, whether renting or owning an accommodation or financial help from family or relatives. Further research should include these cofounders when available. Other limitations come from the design of the study. First, mixed effect models over a four-wave dataset do not track long-term changes over time and, as the nature of the relationship between employment and health may change, estimates found in this study might be different when looking at long-term follow-up. Second, this study has focused on two main definitions of job satisfaction but further studies could look at different specific aspects to distinguish those that play a bigger part in the explanation. Finally, it can be logically assumed that part of the association between SSP and employment is causal (e.g., contract work is, in principle, not a choice) but reverse causation is possible.

Nevertheless, three main findings from this study may be of interest. First, we find that the employment category is associated with SPP. Contract work, that is one of the main features of the Japanese labour market for the workers aged 50 and over, is associated with lower self-predicted retirement age. In comparison with full time workers, we estimate that those in contract work predict an age of death that is about half a year lower. By comparison, retirement also have a negative relationship with SPP but only account for reduction of 0.27 years of age. Second, job satisfaction impacts SPP independently of the employment status but with a much higher effect as those who are not satisfied with their job predict to live 1.3 years less compared with those who are satisfied. Including job satisfaction in the model reduce the relationship with employment category to little indicating the job satisfaction might also translate the position respondents occupy within the labour market. Finally, SPP is highly connected with other measures of health as the inclusion of an adjusted layer controlling for self-reported health, life satisfaction or comorbidity reduces the intensity of the coefficients. However, other health variables reduce the intensity of the coefficients without changing their nature, which indicate that SPP remains a specific dimensions that deserves to be analysed as such.

## Supporting information

Supplementary file 1. Sample selection

Supplementary file 2. Descriptive statistics

Supplementary file 3. Self-predicted life expactancy by employment status (all waves)

Supplementary file 4. Estimates for the mixed effects linear regression and linear quantile mixed effect regression by level of adjustment and MI

Supplementary file 5. Estimates for the mixed effects linear regression and linear quantile mixed effect regression by level of adjustment and MI

Supplementary file 6. Estimates for the mixed effects linear regression and linear quantile mixed effect regression by level of adjustment and MI

Supplementary file 7. Estimates for the mixed effects linear regression and linear quantile mixed effect regression by level of adjustment no MI

Supplementary file 8. Estimates for the mixed effects linear regression and linear quantile mixed effect regression by level of adjustment no MI

Supplementary file 9. Estimates for the mixed effects linear regression and linear quantile mixed effect regression by level of adjustment No MI

## Data Availability

Micro anonymised data from the Japanese Study of Aging and Retirement (JSTAR) is available upon request to the Japan Research Institute of Economy, Trade and Industry (RIETI), https://www.rieti.go.jp/jp/index.html

## Acknowledgement

The Foreign Researcher Invitation Program (FY2019) of the Institute for Labour Policy and Training (JLPT).

## Funding

Jacques Wels is funded by the Belgian National Scientific Fund (FNRS) tenured Associate Research Fellowship (CQ) n° 40010931.

## Conflicts of interest

None declared

## Supplementary material

Supplementary file 1. Sample selection

Supplementary file 2. Descriptive statistics

Supplementary file 3. Self-predicted life expectancy by employment status

Supplementary file 4. Estimates for the mixed effects linear regression and linear quantile mixed effect regression by level of adjustment including multiple imputations - Model 1

Supplementary file 5. Estimates for the mixed effects linear regression and linear quantile mixed effect regression by level of adjustment including multiple imputations – Model 2

Supplementary file 6. Estimates for the mixed effects linear regression and linear quantile mixed effect regression by level of adjustment including multiple imputations – Model 2_bis (sensitivity check)

Supplementary file 7. Estimates for the mixed effects linear regression and linear quantile mixed effect regression by level of adjustment excluding multiple imputations - Model 1

Supplementary file 8. Estimates for the mixed effects linear regression and linear quantile mixed effect regression by level of adjustment excluding multiple imputations – Model 2

Supplementary file 9. Estimates for the mixed effects linear regression and linear quantile mixed effect regression by level of adjustment excluding multiple imputations – Model 2_bis (sensitivity check)

## Notes

### Competing Interest Statement

The authors have declared no competing interest.

### Author Declarations

The study used micro anonymised data from the Japanese Study of Aging and Retirement (JSTAR) that is available upon request to the Japan Research Institute of Economy, Trade and Industry (RIETI), https://www.rieti.go.jp/jp/index.html

