## Supplementary file 1. Sample selection for "Subjective survival probabilities by employment category and job satisfaction among the fifty-plus population in Japan"

**Model 1. Sample composition, refreshment and attrition (complete case for employment participation)**

| **WAVE 1** |  | **WAVE 2** |  | **WAVE 3** |  | **WAVE 4** |  | **Attrition** | |
| --- | --- | --- | --- | --- | --- | --- | --- | --- | --- |
| 2007 |  | 2009 |  |  |  |  |  | Wave 1 to 2 | |
| (5 prefectures) |  | (5 prefectures) |  |  |  |  |  | N= | 1027 |
| N=3,731 |  | N=2,704 |  |  |  |  |  | %= | 27.5 |
|  |  | 2009 |  | 2011 |  |  |  | Wave 2 to 3 | |
|  |  | (2 prefectures) |  | (11 prefectures) |  |  |  | N= | 953 |
|  |  | N=1,400 |  | N=3,151 |  |  |  | %= | 23.2 |
|  |  |  |  | 2011 |  | 2013 |  | Wave 3 to 4 | |
|  |  |  |  | (3 prefectures) |  | (10 prefectures) |  | N= | 1107 |
|  |  |  |  | N=1,951 |  | N=3,995 |  | %= | 21.7 |

**Model 2. Sample composition, refreshment, attrition and drop out (non-working)**

| **WAVE 1** |  | **WAVE 2** |  | **WAVE 3** |  | **WAVE 4** |  | **Attrition and drop out** | |
| --- | --- | --- | --- | --- | --- | --- | --- | --- | --- |
| 2007 |  | 2009 |  |  |  |  |  | Wave 1 to 2 | |
| (5 prefectures) |  | (5 prefectures) |  |  |  |  |  | N= | 732 |
| N=2,151 |  | N=1,419 |  |  |  |  |  | %= | 34.0 |
|  |  | 2009 |  | 2011 |  |  |  | Wave 2 to 3 | |
|  |  | (2 prefectures) |  | (11 prefectures) |  |  |  | N= | 619 |
|  |  | N=686 |  | N= 1,486 |  |  |  | %= | 29.4 |
|  |  |  |  | 2011 |  | 2013 |  | Wave 3 to 4 | |
|  |  |  |  | (3 prefectures) |  | (10 prefectures) |  | N= | 661 |
|  |  |  |  | N=1,061 |  | N=1,886 |  | %= | 26.0 |
