## Supplementary file 2. Descriptive statistics for "Subjective survival probabilities by employment category and job satisfaction among the fifty-plus population in Japan"

| **Wave** |  | **All** | **2007** | **2009** | **2009_2** | **2011** | **2011_3** | **2013** |
| --- | --- | --- | --- | --- | --- | --- | --- | --- |
| N |  | 16932 | 3731 | 2704 | 1400 | 3151 | 1951 | 3995 |
| **SSP** | |  |  |  |  |  |  |  |
|  | NA | 589 | 107 | 149 | 52 | 94 | 72 | 115 |
|  | Mean (SD) | 83.386 (4.407) | 82.649 (4.085) | 82.616 (4.244) | 83.807 (4.478) | 83.883 (4.341) | 82.948 (4.467) | 84.256 (4.573) |
|  | Range | 75.000 - 95.000 | 75.000 - 95.000 | 75.000 - 95.000 | 75.000 - 95.000 | 75.000 - 95.000 | 75.000 - 95.000 | 75.000 - 95.000 |
| **Employment category** | |  |  |  |  |  |  |  |
|  | NA | 155 | 1 | 10 | 2 | 71 | 53 | 18 |
|  | Employed full-time | 2470 (14.7%) | 668 (17.9%) | 389 (14.4%) | 234 (16.7%) | 394 (12.8%) | 347 (18.3%) | 438 (11.0%) |
|  | Company executive | 515 (3.1%) | 136 (3.6%) | 66 (2.4%) | 35 (2.5%) | 87 (2.8%) | 78 (4.1%) | 113 (2.8%) |
|  | Employed contract | 739 (4.4%) | 126 (3.4%) | 116 (4.3%) | 46 (3.3%) | 141 (4.6%) | 114 (6.0%) | 196 (4.9%) |
|  | Employed other | 174 (1.0%) | 36 (1.0%) | 32 (1.2%) | 15 (1.1%) | 21 (0.7%) | 8 (0.4%) | 62 (1.6%) |
|  | Employed part-time | 1931 (11.5%) | 429 (11.5%) | 308 (11.4%) | 158 (11.3%) | 307 (10.0%) | 279 (14.7%) | 450 (11.3%) |
|  | Employed temporary | 89 (0.5%) | 24 (0.6%) | 24 (0.9%) | 4 (0.3%) | 12 (0.4%) | 7 (0.4%) | 18 (0.5%) |
|  | Help in independent business | 736 (4.4%) | 181 (4.9%) | 111 (4.1%) | 33 (2.4%) | 151 (4.9%) | 77 (4.1%) | 183 (4.6%) |
|  | Not retired - keep house | 4198 (25.0%) | 831 (22.3%) | 649 (24.1%) | 349 (25.0%) | 798 (25.9%) | 470 (24.8%) | 1101 (27.7%) |
|  | Not retired - other reasons | 642 (3.8%) | 167 (4.5%) | 103 (3.8%) | 86 (6.2%) | 96 (3.1%) | 94 (5.0%) | 96 (2.4%) |
|  | Not retired - receiving medical care | 624 (3.7%) | 133 (3.6%) | 84 (3.1%) | 74 (5.3%) | 130 (4.2%) | 48 (2.5%) | 155 (3.9%) |
|  | Owner of independent business | 1912 (11.4%) | 513 (13.8%) | 351 (13.0%) | 154 (11.0%) | 347 (11.3%) | 144 (7.6%) | 403 (10.1%) |
|  | Retired | 2624 (15.6%) | 448 (12.0%) | 439 (16.3%) | 203 (14.5%) | 570 (18.5%) | 225 (11.9%) | 739 (18.6%) |
|  | Side job at home | 123 (0.7%) | 38 (1.0%) | 22 (0.8%) | 7 (0.5%) | 26 (0.8%) | 7 (0.4%) | 23 (0.6%) |
| **Job satisfaction (categorical)** | |  |  |  |  |  |  |  |
|  | NA | 8086 | 1580 | 1275 | 712 | 1591 | 838 | 2090 |
|  | Strongly agree | 1573 (17.8%) | 371 (17.2%) | 256 (17.9%) | 141 (20.5%) | 273 (17.5%) | 204 (18.3%) | 328 (17.2%) |
|  | Somewhat agree | 5496 (62.1%) | 1354 (62.9%) | 871 (61.0%) | 385 (56.0%) | 954 (61.2%) | 706 (63.4%) | 1226 (64.4%) |
|  | Do not really agree | 1164 (13.2%) | 289 (13.4%) | 197 (13.8%) | 100 (14.5%) | 233 (14.9%) | 117 (10.5%) | 228 (12.0%) |
|  | Strongly disagree | 613 (6.9%) | 137 (6.4%) | 105 (7.3%) | 62 (9.0%) | 100 (6.4%) | 86 (7.7%) | 123 (6.5%) |
| **Job satisfaction (item based)** | |  |  |  |  |  |  |  |
|  | NA | 9605 | 1917 | 1518 | 872 | 1856 | 1048 | 2394 |
|  | Mean (SD) | 13.929 (2.657) | 13.741 (2.583) | 14.294 (2.635) | 13.453 (2.639) | 14.282 (2.667) | 13.313 (2.638) | 14.088 (2.663) |
|  | Range | 6.143 - 24.571 | 6.143 - 23.429 | 6.143 - 23.429 | 6.143 - 23.571 | 6.143 - 24.571 | 6.143 - 22.429 | 6.143 - 24.429 |
| **Number of days off** | |  |  |  |  |  |  |  |
|  | N-Miss | 10825 | 2063 | 1535 | 903 | 1952 | 1826 | 2546 |
|  | Mean (SD) | 13.670 (29.612) | 24.985 (44.284) | 7.765 (13.538) | 14.272 (30.202) | 7.948 (12.585) | 29.640 (66.250) | 8.558 (14.894) |
|  | Range | 0.000 - 365.000 | 0.000 - 350.000 | 0.000 - 245.000 | 0.000 - 200.000 | 0.000 - 100.000 | 0.000 - 365.000 | 0.000 - 285.000 |
| **Working time** | |  |  |  |  |  |  |  |
|  | NA | 8790 | 1713 | 1408 | 780 | 1740 | 901 | 2248 |
|  | 31 to 40 | 2393 (29.4%) | 536 (26.6%) | 423 (32.6%) | 185 (29.8%) | 411 (29.1%) | 311 (29.6%) | 527 (30.2%) |
|  | 21 to 30 | 1116 (13.7%) | 246 (12.2%) | 165 (12.7%) | 87 (14.0%) | 192 (13.6%) | 148 (14.1%) | 278 (15.9%) |
|  | 41 to 50 | 1785 (21.9%) | 512 (25.4%) | 278 (21.5%) | 151 (24.4%) | 315 (22.3%) | 190 (18.1%) | 339 (19.4%) |
|  | 51 and over | 1164 (14.3%) | 369 (18.3%) | 182 (14.0%) | 103 (16.6%) | 193 (13.7%) | 124 (11.8%) | 193 (11.0%) |
|  | Less than 20 | 1684 (20.7%) | 355 (17.6%) | 248 (19.1%) | 94 (15.2%) | 300 (21.3%) | 277 (26.4%) | 410 (23.5%) |
| **Working time pattern** | | 51.9 |  |  |  |  |  |  |
|  | Not working | 8790 (51.9%) | 1713 (45.9%) | 1408 (52.1%) | 780 (55.7%) | 1740 (55.2%) | 901 (46.2%) | 2248 (56.3%) |
|  | Generally same hours every week, year-round | 6749 (39.9%) | 1630 (43.7%) | 1081 (40.0%) | 524 (37.4%) | 1175 (37.3%) | 863 (44.2%) | 1476 (36.9%) |
|  | Hours vary each week, but I work year-round | 1039 (6.1%) | 278 (7.5%) | 155 (5.7%) | 90 (6.4%) | 168 (5.3%) | 158 (8.1%) | 190 (4.8%) |
|  | Work during some seasons and not others | 354 (2.1%) | 110 (2.9%) | 60 (2.2%) | 6 (0.4%) | 68 (2.2%) | 29 (1.5%) | 81 (2.0%) |
| **Age** | |  |  |  |  |  |  |  |
|  | Mean (SD) | 64.642 (7.663) | 63.359 (7.034) | 65.359 (7.034) | 63.527 (7.258) | 66.870 (7.183) | 62.455 (6.844) | 67.232 |
|  | Range | 43 - 83 | 50 - 77 | 52 - 79 | 51 - 76 | 53 -81 | 47 -75 | 49 - 82 |
| **Gender** | |  |  |  |  |  |  |  |
|  | NA | 1 | 0 | 0 | 0 | 0 | 1 | 0 |
|  | Male | 8301 (49.0%) | 1864 (50.0%) | 1378 (51.0%) | 661 (47.2%) | 1595 (50.6%) | 869 (44.6%) | 1934 (48.4%) |
|  | Female | 8630 (51.0%) | 1867 (50.0%) | 1326 (49.0%) | 739 (52.8%) | 1556 (49.4%) | 1081 (55.4%) | 2061 (51.6%) |
| **Highest education level** | |  |  |  |  |  |  |  |
|  | NA | 86 | 18 | 12 | 8 | 17 | 9 | 22 |
|  | University degree | 2666 (15.8%) | 492 (13.3%) | 337 (12.5%) | 239 (17.2%) | 441 (14.1%) | 467 (24.0%) | 690 (17.4%) |
|  | Elementary to middle school | 4509 (26.8%) | 1195 (32.2%) | 893 (33.2%) | 311 (22.3%) | 934 (29.8%) | 238 (12.3%) | 938 (23.6%) |
|  | High School | 7372 (43.8%) | 1598 (43.0%) | 1143 (42.5%) | 645 (46.3%) | 1366 (43.6%) | 868 (44.7%) | 1752 (44.1%) |
|  | Junior college | 996 (5.9%) | 151 (4.1%) | 105 (3.9%) | 97 (7.0%) | 152 (4.9%) | 213 (11.0%) | 278 (7.0%) |
|  | Vocational school | 1303 (7.7%) | 277 (7.5%) | 214 (7.9%) | 100 (7.2%) | 241 (7.7%) | 156 (8.0%) | 315 (7.9%) |
| **Marital status at baseline** | |  |  |  |  |  |  |  |
|  | NA | 7 | 1 | 2215 (81.9%) | 1 | 1 | 3 | 1 |
|  | Married or common law spouse | 13557 (80.1%) | 3035 (81.4%) | 489 (18.1%) | 1052 (75.2%) | 2537 (80.5%) | 1515 (77.8%) | 3203 (80.2%) |
|  | Not Married and no common law spouse | 3368 (19.9%) | 695 (18.6%) |  | 347 (24.8%) | 613 (19.5%) | 433 (22.2%) | 791 (19.8%) |
| **Expenses** | |  |  |  |  |  |  |  |
|  | NA | 443 | 73 | 127 | 31 | 53 | 46 | 113 |
|  | No | 1818 (11.0%) | 396 (10.8%) | 281 (10.9%) | 164 (12.0%) | 381 (12.3%) | 165 (8.7%) | 431 (11.1%) |
|  | Not known | 112 (0.7%) | 25 (0.7%) | 16 (0.6%) | 10 (0.7%) | 23 (0.7%) | 9 (0.5%) | 29 (0.7%) |
|  | Yes | 14559 (88.3%) | 3237 (88.5%) | 2280 (88.5%) | 1195 (87.3%) | 2694 (87.0%) | 1731 (90.9%) | 3422 (88.2%) |
| **Rent** | |  |  |  |  |  |  |  |
|  | NA | 2991 | 108 | 2532 | 55 | 97 | 76 | 123 |
|  | Own | 11627 (83.4%) | 3142 (86.7%) | 132 (76.7%) | 989 (73.5%) | 2613 (85.6%) | 1465 (78.1%) | 3286 (84.9%) |
|  | Rent | 2262 (16.2%) | 467 (12.9%) | 38 (22.1%) | 346 (25.7%) | 433 (14.2%) | 405 (21.6%) | 573 (14.8%) |
| **Private health insurance** | |  |  |  |  |  |  |  |
|  | NA | 418 | 93 | 56 | 48 | 67 | 65 | 89 |
|  | Mean (SD) | 0.568 (0.495) | 0.561 (0.496) | 0.569 (0.495) | 0.467 (0.499) | 0.545 (0.498) | 0.645 (0.479) | 0.590 (0.492) |
|  | Range | 0.000 - 1.000 | 0.000 - 1.000 | 0.000 - 1.000 | 0.000 - 1.000 | 0.000 - 1.000 | 0.000 - 1.000 | 0.000 - 1.000 |
| **Self-reported health** | |  |  |  |  |  |  |  |
|  | NA | 26 | 7 | 3 | 4 | 1 | 10 | 1 |
|  | Mean (SD) | 2.513 (1.061) | 2.527 (1.099) | 2.515 (1.043) | 2.615 (1.137) | 2.539 (1.007) | 2.436 (1.099) | 2.477 (1.027) |
|  | Range | 1.000 - 5.000 | 1.000 - 5.000 | 1.000 - 5.000 | 1.000 - 5.000 | 1.000 - 5.000 | 1.000 - 5.000 | 1.000 - 5.000 |
| **Multimorbidity** | |  |  |  |  |  |  |  |
|  | NA | 182 | 42 | 57 | 27 | 5 | 26 | 25 |
|  | Mean (SD) | 0.228 (0.420) | 0.246 (0.430) | 0.233 (0.423) | 0.243 (0.429) | 0.203 (0.403) | 0.242 (0.428) | 0.218 (0.413) |
|  | &Range | 0.000 - 1.000 | 0.000 - 1.000 | 0.000 - 1.000 | 0.000 - 1.000 | 0.000 - 1.000 | 0.000 - 1.000 | 0.000 - 1.000 |
| **GHQ casness** | |  |  |  |  |  |  |  |
|  | NA | 2825 | 551 | 493 | 366 | 493 | 322 | 600 |
|  | &Mean (SD) | 0.323 (0.468) | 0.300 (0.458) | 0.311 (0.463) | 0.348 (0.477) | 0.310 (0.463) | 0.317 (0.466) | 0.358 (0.480) |
|  | Range | 0.000 - 1.000 | 0.000 - 1.000 | 0.000 - 1.000 | 0.000 - 1.000 | 0.000 - 1.000 | 0.000 - 1.000 | 0.000 - 1.000 |
| **Outpatient at clinic or hospital** | |  |  |  |  |  |  |  |
|  | NA | 3626 | 46 | 52 | 32 | 1588 | 35 | 1873 |
|  | Mean (SD) | 0.704 (0.457) | 0.690 (0.463) | 0.760 (0.427) | 0.621 (0.485) | 0.666 (0.472) | 0.715 (0.452) | 0.729 (0.445) |
|  | Range | 0.000 - 1.000 | 0.000 - 1.000 | 0.000 - 1.000 | 0.000 - 1.000 | 0.000 - 1.000 | 0.000 - 1.000 | 0.000 - 1.000 |
| **Night at hospital** | |  |  |  |  |  |  |  |
|  | NA | 3597 | 45 | 48 | 28 | 1584 | 30 | 1862 |
|  | Mean (SD) | 0.097 (0.296) | 0.095 (0.293) | 0.098 (0.297) | 0.093 (0.290) | 0.105 (0.307) | 0.078 (0.268) | 0.116 (0.320) |
|  | Range | 0.000 - 1.000 | 0.000 - 1.000 | 0.000 - 1.000 | 0.000 - 1.000 | 0.000 - 1.000 | 0.000 - 1.000 | 0.000 - 1.000 |
| **Life satisfaction** | |  |  |  |  |  |  |  |
|  | NA | 1139 | 124 | 189 | 219 | 185 | 159 | 263 |
|  | Mean (SD) | 0.180 (0.385) | 0.202 (0.401) | 0.183 (0.386) | 0.208 (0.406) | 0.157 (0.364) | 0.204 (0.403) | 0.157 (0.364) |
|  | Range | 0.000 - 1.000 | 0.000 - 1.000 | 0.000 - 1.000 | 0.000 - 1.000 | 0.000 - 1.000 | 0.000 - 1.000 | 0.000 - 1.000 |
