## Supplementary figures and images for "Subjective survival probabilities by employment category and job satisfaction among the fifty-plus population in Japan"

### Supplementary file 3. Self-predicted life expactancy by employment status (all waves)

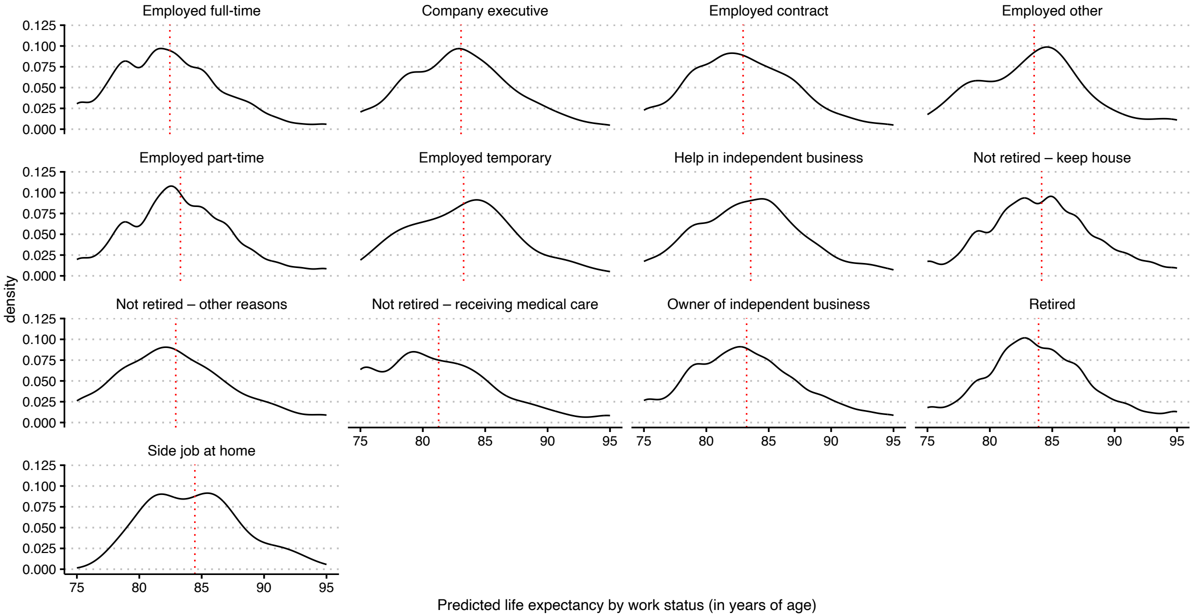
