## Supplementary file 4. Estimates for the mixed effects linear regression and linear quantile mixed effect regression by level of adjustment and MI for "Subjective survival probabilities by employment category and job satisfaction among the fifty-plus population in Japan"

**Supplementary file X. Estimates for the mixed effects linear regression and linear quantile mixed effect regression by level of adjustment including multiple imputations – Model 1**

|  |  | **Multiple imputations** | | | | | | |
| --- | --- | --- | --- | --- | --- | --- | --- | --- |
|  |  | **Mixed Effects Linear Regression** | | |  | **Linear Quantile Mixed Effect Regression** | | |
|  |  | Unadjusted | Socio-demographic adjustment | Health adjustment |  | Unadjusted | Socio-demographic adjustment | Health adjustment |
|  | (Intercept) | 75.092^*^ | 74.971^*^ | 76.004^*^ |  | 74.501^*^ | 74.386^*^ | 75.576^*^ |
|  |  | [74.302; 75.881] | [74.066; 75.875] | [75.071; 76.938] |  | [73.852; 75.149] | [73.586; 75.185] | [74.731; 76.422] |
| Employment | Company executive | -0.12 | -0.146 | -0.16 |  | -0.148 | -0.18 | -0.196 |
|  |  | [-0.472; 0.233] | [-0.496; 0.204] | [-0.521; 0.201] |  | [-0.544; 0.247] | [-0.586; 0.226] | [-0.596; 0.205] |
|  | Employed contract | -0.400^*^ | -0.402^*^ | -0.419^*^ |  | -0.503^*^ | -0.499^*^ | -0.511^*^ |
|  |  | [-0.689; -0.112] | [-0.692; -0.111] | [-0.703; -0.135] |  | [-0.776; -0.229] | [-0.798; -0.201] | [-0.786; -0.235] |
|  | Employed other | -0.143 | -0.147 | -0.171 |  | -0.183 | -0.192 | -0.216 |
|  |  | [-0.810; 0.524] | [-0.821; 0.527] | [-0.832; 0.490] |  | [-0.841; 0.476] | [-0.851; 0.468] | [-0.910; 0.478] |
|  | Employed part-time | -0.067 | -0.053 | -0.064 |  | -0.137 | -0.123 | -0.135 |
|  |  | [-0.342; 0.209] | [-0.332; 0.226] | [-0.331; 0.203] |  | [-0.410; 0.136] | [-0.408; 0.161] | [-0.405; 0.134] |
|  | Employed temporary | -0.036 | -0.023 | -0.009 |  | -0.213 | -0.195 | -0.159 |
|  |  | [-0.900; 0.827] | [-0.901; 0.854] | [-0.846; 0.828] |  | [-1.157; 0.731] | [-1.118; 0.728] | [-1.078; 0.759] |
|  | Help in independent business | -0.076 | -0.086 | -0.086 |  | -0.116 | -0.133 | -0.127 |
|  |  | [-0.382; 0.231] | [-0.394; 0.222] | [-0.394; 0.222] |  | [-0.432; 0.200] | [-0.432; 0.165] | [-0.459; 0.205] |
|  | House keeping | 0.035 | 0.026 | 0.069 |  | -0.049 | -0.063 | 0.018 |
|  |  | [-0.191; 0.261] | [-0.200; 0.251] | [-0.149; 0.287] |  | [-0.275; 0.177] | [-0.296; 0.169] | [-0.196; 0.231] |
|  | Not retired – other reasons | -0.211 | -0.187 | -0.081 |  | -0.335 | -0.299 | -0.156 |
|  |  | [-0.611; 0.190] | [-0.585; 0.211] | [-0.472; 0.310] |  | [-0.757; 0.087] | [-0.705; 0.107] | [-0.552; 0.239] |
|  | Receiving medical care | -1.348^*^ | -1.319^*^ | -0.825^*^ |  | -1.595^*^ | -1.546^*^ | -0.918^*^ |
|  |  | [-1.760; -0.936] | [-1.724; -0.914] | [-1.227; -0.424] |  | [-2.046; -1.145] | [-1.986; -1.105] | [-1.345; -0.491] |
|  | Owner of independent business | -0.079 | -0.079 | -0.088 |  | -0.153 | -0.15 | -0.159 |
|  |  | [-0.378; 0.220] | [-0.379; 0.222] | [-0.371; 0.195] |  | [-0.465; 0.159] | [-0.456; 0.157] | [-0.457; 0.139] |
|  | Retired | -0.306^*^ | -0.316^*^ | -0.248^*^ |  | -0.358^*^ | -0.378^*^ | -0.272^*^ |
|  |  | [-0.528; -0.085] | [-0.540; -0.092] | [-0.475; -0.021] |  | [-0.590; -0.127] | [-0.615; -0.140] | [-0.512; -0.032] |
|  | Side job at home | 0.337 | 0.348 | 0.3 |  | 0.324 | 0.34 | 0.278 |
|  |  | [-0.348; 1.023] | [-0.336; 1.031] | [-0.377; 0.977] |  | [-0.399; 1.047] | [-0.382; 1.062] | [-0.424; 0.980] |
|  | age | 0.126^*^ | 0.130^*^ | 0.134^*^ |  | 0.126^*^ | 0.130^*^ | 0.135^*^ |
|  |  | [ 0.115; 0.136] | [ 0.117; 0.142] | [ 0.121; 0.147] |  | [ 0.115; 0.137] | [ 0.117; 0.143] | [ 0.121; 0.148] |
|  | gender: Female | 0.403^*^ | 0.453^*^ | 0.433^*^ |  | 0.425^*^ | 0.477^*^ | 0.458^*^ |
|  |  | [ 0.268; 0.539] | [ 0.315; 0.591] | [ 0.296; 0.570] |  | [ 0.279; 0.571] | [ 0.336; 0.618] | [ 0.327; 0.589] |
| Education | Elementary to middle school | | -0.280^*^ | -0.237 |  |  | -0.269^*^ | -0.21 |
|  |  |  | [-0.528; -0.033] | [-0.477; 0.004] |  |  | [-0.529; -0.010] | [-0.459; 0.039] |
|  | High School |  | -0.208 | -0.183 |  |  | -0.225^*^ | -0.210^*^ |
|  |  |  | [-0.424; 0.008] | [-0.390; 0.025] |  |  | [-0.427; -0.024] | [-0.412; -0.008] |
|  | Juniorcollege |  | 0.06 | 0.038 |  |  | 0.059 | 0.037 |
|  |  |  | [-0.338; 0.458] | [-0.365; 0.441] |  |  | [-0.348; 0.465] | [-0.361; 0.435] |
|  | Vocationalschool |  | -0.298^*^ | -0.262 |  |  | -0.306^*^ | -0.262 |
|  |  |  | [-0.582; -0.013] | [-0.538; 0.014] |  |  | [-0.601; -0.011] | [-0.557; 0.034] |
|  | University degree |  | Ref. | Ref. |  |  | Ref. | Ref. |
| Marital Status | Not Married and no common law spouse | | -0.135 | -0.038 |  |  | -0.144 | -0.03 |
|  |  |  | [-0.327; 0.057] | [-0.224; 0.148] |  |  | [-0.330; 0.042] | [-0.202; 0.143] |
|  | Maried or common law spouse |  | Ref. | Ref. |  |  | Ref. | Ref. |
| Ask friends or family to cover expenses | Yes |  | 0.134 | 0.088 |  |  | 0.142 | 0.071 |
|  |  |  | [-0.061; 0.329] | [-0.103; 0.279] |  |  | [-0.055; 0.339] | [-0.131; 0.273] |
|  | No |  | Ref. | Ref. |  |  | Ref. | Ref. |
| Rent accomodation | Yes |  | -0.174^*^ | -0.028 |  |  | -0.238^*^ | -0.054 |
|  |  |  | [-0.333; -0.016] | [-0.184; 0.127] |  |  | [-0.415; -0.061] | [-0.222; 0.114] |
|  | No |  | Ref. | Ref. |  |  | Ref. | Ref. |
| Private health insurance | Yes |  | -0.043 | -0.106 |  |  | -0.061 | -0.14 |
|  |  |  | [-0.194; 0.108] | [-0.262; 0.049] |  |  | [-0.215; 0.093] | [-0.301; 0.020] |
|  | No |  | Ref. | Ref. |  |  | Ref. | Ref. |
| Self-reported health | |  |  | -0.367^*^ |  |  |  | -0.399^*^ |
|  |  |  |  | [-0.416; -0.318] |  |  |  | [-0.462; -0.336] |
| Multimorbidity | Yes |  |  | -0.348^*^ |  |  |  | -0.350^*^ |
|  |  |  |  | [-0.503; -0.194] |  |  |  | [-0.504; -0.195] |
|  | No |  |  | Ref. |  |  |  | Ref. |
| GHQ casness | Yes |  |  | -0.291^*^ |  |  |  | -0.296^*^ |
|  |  |  |  | [-0.436; -0.147] |  |  |  | [-0.459; -0.133] |
|  | No |  |  | Ref. |  |  |  | Ref. |
| Outpatient at clinic or hospital | Yes |  |  | -0.131 |  |  |  | -0.143^*^ |
|  |  |  |  | [-0.265; 0.002] |  |  |  | [-0.282; -0.005] |
|  | No |  |  | Ref. |  |  |  | Ref. |
| Night at hospital | Yes |  |  | -0.315^*^ |  |  |  | -0.301^*^ |
|  |  |  |  | [-0.559; -0.072] |  |  |  | [-0.540; -0.062] |
|  | No |  |  | Ref. |  |  |  | Ref. |
| Life satisfaction | Yes |  |  | -0.724^*^ |  |  |  | -0.789^*^ |
|  |  |  |  | [-0.937; -0.510] |  |  |  | [-0.993; -0.585] |
|  | No |  |  | Ref. |  |  |  | Ref. |
| Num. obs. | | 16197 | 13364 | 8448 |  | 16197 | 13364 | 8448 |
| Num. groups: id | | 6951 | 6725 | 5830 |  | 6951 | 6725 | 5830 |
| Num. groups: time | | 4 | 4 | 4 |  | 4 | 4 | 4 |

* Null hypothesis value outside the 95% confidence interval.
