## Supplementary file 5. Estimates for the mixed effects linear regression and linear quantile mixed effect regression by level of adjustment and MI for "Subjective survival probabilities by employment category and job satisfaction among the fifty-plus population in Japan"

**Supplementary file X. Estimates for the mixed effects linear regression and linear quantile mixed effect regression by level of adjustment including multiple imputations – Model 2**

|  |  | **Multiple imputations** | | | | | | |
| --- | --- | --- | --- | --- | --- | --- | --- | --- |
|  |  | **Mixed Effects Linear Regression** | | |  | **Linear Quantile Mixed Effect Regression** | | |
|  |  | Unadjusted | Socio-demographic adjustment | Health adjustment |  | Unadjusted | Socio-demographic adjustment | Health adjustment |
|  | (Intercept) | 75.163^*^ | 74.852^*^ | 75.763^*^ |  | 74.801^*^ | 74.516^*^ | 75.715^*^ |
|  |  | [73.843; 76.483] | [73.429; 76.275] | [74.342; 77.185] |  | [73.385; 76.217] | [72.994; 76.038] | [74.354; 77.075] |
| Employment | Company executive | -0.165 | -0.222 | -0.243 |  | -0.073 | -0.147 | -0.138 |
|  |  | [-0.571; 0.242] | [-0.632; 0.188] | [-0.650; 0.164] |  | [-0.548; 0.402] | [-0.579; 0.285] | [-0.597; 0.321] |
|  | Employed contract | -0.124 | -0.131 | -0.155 |  | -0.18 | -0.185 | -0.195 |
|  |  | [-0.446; 0.199] | [-0.454; 0.192] | [-0.474; 0.165] |  | [-0.520; 0.161] | [-0.540; 0.170] | [-0.567; 0.176] |
|  | Employed other | 0.103 | 0.088 | 0.054 |  | 0.011 | -0.033 | -0.064 |
|  |  | [-0.517; 0.722] | [-0.531; 0.707] | [-0.562; 0.670] |  | [-0.634; 0.657] | [-0.791; 0.724] | [-0.767; 0.639] |
|  | Employed temporary | 0.393 | 0.398 | 0.346 |  | 0.331 | 0.315 | 0.263 |
|  |  | [-0.491; 1.276] | [-0.487; 1.283] | [-0.531; 1.223] |  | [-0.735; 1.396] | [-0.731; 1.360] | [-0.700; 1.227] |
|  | Help in independent business | -0.118 | -0.149 | -0.108 |  | -0.113 | -0.154 | -0.072 |
|  |  | [-0.492; 0.257] | [-0.524; 0.227] | [-0.479; 0.263] |  | [-0.524; 0.298] | [-0.565; 0.257] | [-0.477; 0.333] |
|  | Owner of independent business | -0.02 | -0.035 | -0.02 |  | 0.027 | 0.023 | 0.024 |
|  |  | [-0.302; 0.262] | [-0.321; 0.251] | [-0.303; 0.264] |  | [-0.308; 0.362] | [-0.294; 0.339] | [-0.283; 0.330] |
|  | Side job at home | 0.492 | 0.494 | 0.49 |  | 0.707 | 0.716 | 0.661 |
|  |  | [-0.342; 1.325] | [-0.343; 1.330] | [-0.336; 1.316] |  | [-0.256; 1.671] | [-0.208; 1.640] | [-0.184; 1.506] |
|  | Job satisfaction: somewhat satisfied | -0.206 | -0.188 | -0.108 |  | -0.283^*^ | -0.252 | -0.077 |
|  |  | [-0.441; 0.029] | [-0.422; 0.046] | [-0.342; 0.126] |  | [-0.543; -0.023] | [-0.526; 0.021] | [-0.347; 0.194] |
|  | Job satisfaction: not really satisfied | -0.608^*^ | -0.568^*^ | -0.354^*^ |  | -0.746^*^ | -0.707^*^ | -0.317 |
|  |  | [-0.927; -0.289] | [-0.888; -0.248] | [-0.674; -0.034] |  | [-1.099; -0.393] | [-1.067; -0.348] | [-0.674; 0.040] |
|  | Job satisfaction: strongly not satisfied | -1.495^*^ | -1.455^*^ | -1.211^*^ |  | -1.872^*^ | -1.831^*^ | -1.373^*^ |
|  |  | [-1.904; -1.086] | [-1.870; -1.040] | [-1.622; -0.799] |  | [-2.332; -1.411] | [-2.316; -1.346] | [-1.842; -0.905] |
|  | Days off | -0.002 | -0.002 | -0.001 |  | -0.002 | -0.003 | 0 |
|  |  | [-0.005; 0.002] | [-0.005; 0.002] | [-0.005; 0.002] |  | [-0.006; 0.003] | [-0.007; 0.002] | [-0.004; 0.003] |
|  | Working time 21 to 30h | -0.022 | -0.019 | 0.027 |  | -0.094 | -0.116 | -0.043 |
|  |  | [-0.313; 0.270] | [-0.310; 0.271] | [-0.261; 0.316] |  | [-0.428; 0.240] | [-0.429; 0.197] | [-0.377; 0.291] |
|  | Working time 41 to 50h | 0.181 | 0.185 | 0.176 |  | 0.099 | 0.082 | 0.068 |
|  |  | [-0.079; 0.441] | [-0.078; 0.447] | [-0.087; 0.439] |  | [-0.189; 0.387] | [-0.229; 0.392] | [-0.217; 0.354] |
|  | Working time 51h and over | 0.3 | 0.309^*^ | 0.283 |  | 0.326 | 0.348 | 0.311 |
|  |  | [-0.002; 0.602] | [ 0.009; 0.609] | [-0.016; 0.583] |  | [-0.003; 0.656] | [-0.009; 0.704] | [-0.054; 0.676] |
|  | Working time Less than 20h | 0.129 | 0.121 | 0.155 |  | 0.056 | 0.055 | 0.121 |
|  |  | [-0.162; 0.420] | [-0.172; 0.414] | [-0.136; 0.445] |  | [-0.250; 0.362] | [-0.248; 0.358] | [-0.230; 0.472] |
|  | Working time pattern: Hours vary each week, but I work year-round | 0.234 | 0.224 | 0.24 |  | 0.203 | 0.22 | 0.254 |
|  |  | [-0.167; 0.634] | [-0.178; 0.626] | [-0.162; 0.642] |  | [-0.275; 0.681] | [-0.246; 0.686] | [-0.191; 0.700] |
|  | Working time pattern: I work during some seasons and not others | -0.09 | -0.1 | -0.06 |  | -0.095 | -0.09 | 0.004 |
|  |  | [-0.622; 0.442] | [-0.634; 0.433] | [-0.594; 0.474] |  | [-0.628; 0.438] | [-0.649; 0.470] | [-0.508; 0.516] |
|  | age | 0.120^*^ | 0.125^*^ | 0.131^*^ |  | 0.119^*^ | 0.124^*^ | 0.128^*^ |
|  |  | [ 0.101; 0.139] | [ 0.104; 0.145] | [ 0.111; 0.151] |  | [ 0.099; 0.139] | [ 0.102; 0.147] | [ 0.106; 0.150] |
|  | gender: Female | 0.836^*^ | 0.892^*^ | 0.826^*^ |  | 0.820^*^ | 0.860^*^ | 0.784^*^ |
|  |  | [ 0.584; 1.089] | [ 0.622; 1.161] | [ 0.561; 1.090] |  | [ 0.560; 1.080] | [ 0.568; 1.151] | [ 0.505; 1.064] |
| Education | Elementary to middle school | | -0.271 | -0.286 |  |  | -0.227 | -0.237 |
|  |  |  | [-0.660; 0.117] | [-0.666; 0.095] |  |  | [-0.639; 0.185] | [-0.630; 0.155] |
|  | High School |  | -0.298 | -0.277 |  |  | -0.228 | -0.227 |
|  |  |  | [-0.620; 0.025] | [-0.590; 0.037] |  |  | [-0.565; 0.110] | [-0.554; 0.100] |
|  | Juniorcollege |  | 0.226 | 0.187 |  |  | 0.255 | 0.19 |
|  |  |  | [-0.313; 0.765] | [-0.341; 0.714] |  |  | [-0.312; 0.822] | [-0.350; 0.729] |
|  | Vocationalschool |  | -0.356 | -0.303 |  |  | -0.342 | -0.275 |
|  |  |  | [-0.841; 0.130] | [-0.778; 0.171] |  |  | [-0.855; 0.170] | [-0.773; 0.224] |
|  | University degree |  | Ref. | Ref. |  |  | Ref. | Ref. |
| Marital Status | Not Married and no common law spouse | | -0.081 | 0.017 |  |  | -0.028 | 0.075 |
|  |  |  | [-0.398; 0.235] | [-0.292; 0.326] |  |  | [-0.358; 0.302] | [-0.250; 0.401] |
|  | Married or common law spouse |  | Ref. | Ref. |  |  | Ref. | Ref. |
| Ask friends or family to cover expenses | Yes |  | -0.381 | -0.398 |  |  | -0.285 | -0.327 |
|  |  |  | [-1.381; 0.618] | [-1.385; 0.588] |  |  | [-1.290; 0.719] | [-1.460; 0.805] |
|  | No |  | Ref. | Ref. |  |  | Ref. | Ref. |
| Rent accommodation | Yes |  | [-1.878; 0.998] | [-1.738; 1.074] |  |  | [-1.486; 1.502] | [-1.404; 1.690] |
|  |  |  | -0.088 | -0.009 |  |  | -0.127 | 0.008 |
|  | No |  | Ref. | Ref. |  |  | Ref. | Ref. |
| Private health insurance | Yes |  | -0.09 | -0.098 |  |  | -0.137 | -0.162 |
|  |  |  | [-0.348; 0.168] | [-0.353; 0.156] |  |  | [-0.398; 0.125] | [-0.394; 0.071] |
|  | No |  | Ref. | Ref. |  |  | Ref. | Ref. |
| Self-reported health | |  |  | -0.425^*^ |  |  |  | -0.469^*^ |
|  |  |  |  | [-0.517; -0.332] |  |  |  | [-0.574; -0.363] |
| Multimorbidity | Yes |  |  | -0.498^*^ |  |  |  | -0.503^*^ |
|  |  |  |  | [-0.749; -0.247] |  |  |  | [-0.784; -0.221] |
|  | No |  |  | Ref. |  |  |  | Ref. |
| GHQ caseness | Yes |  |  | -0.197 |  |  |  | -0.208 |
|  |  |  |  | [-0.425; 0.032] |  |  |  | [-0.440; 0.024] |
|  | No |  |  | Ref. |  |  |  | Ref. |
| Outpatient at clinic or hospital | Yes |  |  | -0.266^*^ |  |  |  | -0.305^*^ |
|  |  |  |  | [-0.471; -0.062] |  |  |  | [-0.516; -0.094] |
|  | No |  |  | Ref. |  |  |  | Ref. |
| Night at hospital | Yes |  |  | -0.304 |  |  |  | -0.34 |
|  |  |  |  | [-0.638; 0.029] |  |  |  | [-0.701; 0.021] |
|  | No |  |  | Ref. |  |  |  | Ref. |
| Life satisfaction | Yes |  |  | -0.278^*^ |  |  |  | -0.454^*^ |
|  |  |  |  | [-0.544; -0.011] |  |  |  | [-0.746; -0.163] |
|  | No |  |  | Ref. |  |  |  | Ref. |
| Num. obs. | | 5671 | 4549 | 3130 |  | 5671 | 4549 | 3130 |
| Num. groups: id | | 3079 | 2868 | 2263 |  | 3079 | 2868 | 2263 |
| Num. groups: time | | 4 | 4 | 4 |  | 4 | 4 | 4 |

* Null hypothesis value outside the 95% confidence interval.
