## Supplementary file 8. Estimates for the mixed effects linear regression and linear quantile mixed effect regression by level of adjustment no MI for "Subjective survival probabilities by employment category and job satisfaction among the fifty-plus population in Japan"

**Supplementary file X. Estimates for the mixed effects linear regression and linear quantile mixed effect regression by level of adjustment excluding multiple imputations – Model 2**

|  |  | **Missing data excluded** | | | | | | |
| --- | --- | --- | --- | --- | --- | --- | --- | --- |
|  |  | **Mixed Effects Linear Regression** | | |  | **Linear Quantile Mixed Effect Regression** | | |
|  |  | Unadjusted | Socio-demographic adjustment | Health adjustment |  | Unadjusted | Socio-demographic adjustment | Health adjustment |
|  | (Intercept) | 75.613^*^ | 74.689^*^ | 76.771^*^ |  | 75.308^*^ | 74.206^*^ | 76.697^*^ |
|  |  | [74.109; 77.117] | [72.990; 76.388] | [74.737; 78.805] |  | [73.684; 76.933] | [72.718; 75.694] | [74.728; 78.666] |
| Employment | Company executive | 0.055 | -0.167 | -0.018 |  | 0.016 | -0.155 | 0.111 |
|  |  | [-0.441; 0.550] | [-0.713; 0.379] | [-0.634; 0.598] |  | [-0.515; 0.548] | [-0.820; 0.509] | [-0.495; 0.717] |
|  | Employed contract | -0.143 | -0.168 | 0.09 |  | -0.172 | -0.189 | 0.01 |
|  |  | [-0.523; 0.238] | [-0.601; 0.264] | [-0.410; 0.590] |  | [-0.607; 0.263] | [-0.674; 0.297] | [-0.604; 0.624] |
|  | Employed other | 0.364 | 0.183 | -0.286 |  | 0.276 | 0.081 | -0.349 |
|  |  | [-0.407; 1.134] | [-0.696; 1.062] | [-1.466; 0.894] |  | [-0.568; 1.119] | [-0.925; 1.087] | [-1.618; 0.921] |
|  | Employed temporary | 0.734 | 1.456^*^ | 1.403 |  | 0.653 | 1.274 | 1.615 |
|  |  | [-0.294; 1.762] | [ 0.177; 2.736] | [-0.180; 2.986] |  | [-0.758; 2.063] | [-0.383; 2.932] | [-0.190; 3.419] |
|  | Help in independent business | -0.012 | -0.302 | -0.376 |  | -0.064 | -0.369 | -0.289 |
|  |  | [-0.493; 0.469] | [-0.836; 0.233] | [-1.099; 0.346] |  | [-0.666; 0.537] | [-0.938; 0.200] | [-0.989; 0.411] |
|  | Owner of independent business | 0.204 | 0.317 | 0.536^*^ |  | 0.19 | 0.296 | 0.591^*^ |
|  |  | [-0.137; 0.545] | [-0.056; 0.690] | [ 0.076; 0.996] |  | [-0.226; 0.607] | [-0.164; 0.757] | [ 0.079; 1.103] |
|  | Side job at home | -0.145 | -0.682 | -0.428 |  | 0.152 | -0.531 | -0.5 |
|  |  | [-1.364; 1.074] | [-2.013; 0.650] | [-2.352; 1.497] |  | [-0.968; 1.273] | [-1.588; 0.527] | [-2.292; 1.293] |
|  | satis2 | -0.232 | -0.2 | -0.155 |  | -0.296 | -0.322 | -0.216 |
|  |  | [-0.505; 0.041] | [-0.512; 0.113] | [-0.532; 0.221] |  | [-0.599; 0.008] | [-0.672; 0.027] | [-0.601; 0.169] |
|  | satis3 | -0.702^*^ | -0.716^*^ | -0.619^*^ |  | -0.823^*^ | -0.851^*^ | -0.521 |
|  |  | [-1.075; -0.328] | [-1.143; -0.288] | [-1.135; -0.103] |  | [-1.258; -0.388] | [-1.325; -0.378] | [-1.068; 0.026] |
|  | satis4 | -1.370^*^ | -1.391^*^ | -1.077^*^ |  | -1.690^*^ | -1.707^*^ | -1.368^*^ |
|  |  | [-1.868; -0.872] | [-1.969; -0.814] | [-1.798; -0.355] |  | [-2.228; -1.152] | [-2.343; -1.071] | [-2.022; -0.713] |
|  | daysoff | -0.001 | -0.002 | -0.003 |  | -0.002 | -0.003 | -0.003 |
|  |  | [-0.005; 0.002] | [-0.006; 0.001] | [-0.007; 0.002] |  | [-0.005; 0.002] | [-0.009; 0.003] | [-0.009; 0.003] |
|  | Workingtime 21 to 30h | -0.301 | -0.462^*^ | -0.494 |  | -0.381 | -0.529^*^ | -0.476 |
|  |  | [-0.655; 0.054] | [-0.862; -0.062] | [-0.994; 0.006] |  | [-0.787; 0.026] | [-0.910; -0.148] | [-0.954; 0.002] |
|  | Workingtime 41 to 50h | 0.272 | 0.224 | 0.229 |  | 0.185 | 0.184 | 0.116 |
|  |  | [-0.008; 0.552] | [-0.093; 0.541] | [-0.140; 0.597] |  | [-0.129; 0.499] | [-0.182; 0.550] | [-0.266; 0.498] |
|  | Workingtime 51h and over | 0.272 | 0.264 | 0.131 |  | 0.355 | 0.294 | 0.035 |
|  |  | [-0.075; 0.619] | [-0.127; 0.654] | [-0.326; 0.588] |  | [-0.072; 0.782] | [-0.154; 0.743] | [-0.408; 0.477] |
|  | Workingtime Less than 20h | 0.046 | -0.012 | -0.081 |  | -0.075 | -0.117 | -0.093 |
|  |  | [-0.303; 0.395] | [-0.405; 0.381] | [-0.586; 0.424] |  | [-0.464; 0.314] | [-0.546; 0.313] | [-0.676; 0.490] |
|  | Working time pattern: Hours vary each week, but I work year-round | -0.083 | -0.035 | 0.23 |  | -0.173 | -0.16 | 0.106 |
|  |  | [-0.407; 0.241] | [-0.402; 0.333] | [-0.230; 0.690] |  | [-0.568; 0.223] | [-0.571; 0.251] | [-0.362; 0.573] |
|  | Working time pattern: I work during some seasons and not others | 0.393 | -0.007 | 3.25 |  | 0.076 | -0.127 | 3.149^*^ |
|  |  | [-0.736; 1.522] | [-4.751; 4.738] | [-4.839; 11.339] |  | [-1.079; 1.230] | [-3.242; 2.988] | [ 1.902; 4.397] |
|  | age | 0.119^*^ | 0.135^*^ | 0.126^*^ |  | 0.116^*^ | 0.135^*^ | 0.120^*^ |
|  |  | [ 0.096; 0.142] | [ 0.109; 0.162] | [ 0.095; 0.157] |  | [ 0.088; 0.144] | [ 0.111; 0.159] | [ 0.087; 0.153] |
|  | gender: Female | 0.995^*^ | 1.182^*^ | 1.111^*^ |  | 1.017^*^ | 1.191^*^ | 1.080^*^ |
|  |  | [ 0.695; 1.296] | [ 0.844; 1.521] | [ 0.716; 1.506] |  | [ 0.704; 1.329] | [ 0.777; 1.605] | [ 0.646; 1.514] |
| Education | Elementary to middle school | | -0.561^*^ | -0.435 |  |  | -0.590^*^ | -0.387 |
|  |  |  | [-1.043; -0.080] | [-0.988; 0.117] |  |  | [-1.107; -0.072] | [-0.872; 0.097] |
|  | High School |  | -0.378 | -0.193 |  |  | -0.375 | -0.216 |
|  |  |  | [-0.770; 0.014] | [-0.622; 0.236] |  |  | [-0.810; 0.060] | [-0.733; 0.302] |
|  | Juniorcollege |  | 0.036 | 0.087 |  |  | 0.125 | 0.147 |
|  |  |  | [-0.634; 0.705] | [-0.664; 0.838] |  |  | [-0.479; 0.729] | [-0.593; 0.887] |
|  | Vocationalschool |  | -0.379 | -0.088 |  |  | -0.324 | -0.073 |
|  |  |  | [-0.955; 0.197] | [-0.726; 0.551] |  |  | [-1.025; 0.377] | [-0.772; 0.626] |
|  | University degree |  | Ref. | Ref. |  |  | Ref. | Ref. |
| Marital Status | Not Married and no common law spouse | | -0.104 | -0.032 |  |  | -0.122 | -0.042 |
|  |  |  | [-0.490; 0.283] | [-0.482; 0.418] |  |  | [-0.566; 0.323] | [-0.542; 0.459] |
|  | Maried or common law spouse |  | Ref. | Ref. |  |  | Ref. | Ref. |
| Ask friends or family to cover expensesYes | Yes |  | 0.458^*^ | 0.481 |  |  | 0.444^*^ | 0.477^*^ |
|  |  |  | [ 0.065; 0.850] | [-0.001; 0.963] |  |  | [ 0.096; 0.792] | [ 0.005; 0.950] |
|  | No |  | Ref. | Ref. |  |  | Ref. | Ref. |
| Rent accomodation | Yes |  | 0.204 | 0.278 |  |  | 0.176 | 0.402 |
|  |  |  | [-0.179; 0.586] | [-0.168; 0.723] |  |  | [-0.234; 0.586] | [-0.105; 0.910] |
|  | No |  | Ref. | Ref. |  |  | Ref. | Ref. |
| Private health insurance | Yes |  | -0.078 | -0.139 |  |  | -0.101 | -0.159 |
|  |  |  | [-0.385; 0.229] | [-0.492; 0.215] |  |  | [-0.373; 0.171] | [-0.505; 0.187] |
|  | No |  | Ref. | Ref. |  |  | Ref. | Ref. |
| Self-reported health | |  |  | -0.457^*^ |  |  |  | -0.546^*^ |
|  |  |  |  | [-0.611; -0.302] |  |  |  | [-0.728; -0.365] |
| Multimorbidity | Yes |  |  | -0.509^*^ |  |  |  | -0.517^*^ |
|  |  |  |  | [-0.903; -0.114] |  |  |  | [-0.871; -0.162] |
|  | No |  |  | Ref. |  |  |  | Ref. |
| GHQ casness | Yes |  |  | -0.283 |  |  |  | -0.276 |
|  |  |  |  | [-0.603; 0.037] |  |  |  | [-0.678; 0.125] |
|  | No |  |  | Ref. |  |  |  | Ref. |
| Outpatient at clinic or hospital | Yes |  |  | -0.571^*^ |  |  |  | -0.685^*^ |
|  |  |  |  | [-0.874; -0.268] |  |  |  | [-0.980; -0.390] |
|  | No |  |  | Ref. |  |  |  | Ref. |
| Night at hospital | Yes |  |  | -0.467 |  |  |  | -0.527 |
|  |  |  |  | [-1.002; 0.068] |  |  |  | [-1.074; 0.021] |
|  | No |  |  | Ref. |  |  |  | Ref. |
| Life satisfaction | Yes |  |  | -0.27 |  |  |  | -0.428^*^ |
|  |  |  |  | [-0.660; 0.120] |  |  |  | [-0.811; -0.045] |
|  | No |  |  | Ref. |  |  |  | Ref. |
| Num. obs. | | 5671 | 4549 | 3130 |  | 5671 | 4549 | 3130 |
| Num. groups: id | | 3079 | 2868 | 2263 |  | 3079 | 2868 | 2263 |
| Num. groups: time | | 4 | 4 | 4 |  | 4 | 4 | 4 |

* Null hypothesis value outside the 95% confidence interval.
